# Prevalence and causes of blindness and visual impairment in Kogi state, Nigeria – Findings from a Rapid Assessment of Avoidable Blindness survey

**DOI:** 10.1101/2023.11.01.23297918

**Authors:** Selben Penzin, Emma Jolley, Kolawole Ogundimu, Caleb Mpyet, Nazaradden Ibrahim, Foluso Owoeye, Sunday Isiyaku, Joy Shu’aibu, Elena Schmidt

## Abstract

**Purpose:** To determine the prevalence and causes of blindness and visual impairment among adults in Kogi, Nigeria.

**Methods:** A Rapid assessment of avoidable blindness (RAAB) protocol was used with additional tools measuring disability and household wealth to measure the prevalence of blindness and visual impairment (VI) and associations with sex, disability, wealth, cataract surgical coverage and its effectiveness.

**Results:** Age- and sex-adjusted all-cause prevalence of bilateral blindness was 3.6% (95%CI 3.0-4.2%), prevalence of blindness among people living with additional, non-visual disabilities was 38.3% (95% CI 29.0-48.6%) compared to 1.6% (95%CI 1.2-2.1%; 𝜒^2^ = 771.9, p<0.001) among people without additional disabilities. Cataract was the principal cause of bilateral blindness (55.3%). Cataract surgical coverage (CSC) at visual acuity (VA) 3/60 was 48.0%, higher among men than women (53.7% vs 40.3%); 12.0% among people with non-visual disabilities; 66.9% among people without non-visual disabilities, higher among people in the wealthiest two quintiles (41.1%) compared to the lowest three (24.3%).

Effective Cataract Surgical Coverage at Visual Acuity 6/60 was 31.0%, higher among males (34.9%) than females (25.5%), low among people with additional, non-visual disabilities (1.9%) compared to people with no additional disabilities (46.2%), higher among people in the wealthiest two quintiles (411%) compared to the poorest three (24.3%).

Good surgical outcome (VA>6/18) was seen in 61 eyes (52.6%) increasing to 71 (61.2%) eyes with best correction. Cost was identified as the main barrier to surgery.

**Conclusion:** Findings suggest there exists inequalities in eye care with women, poorer people and people with disabilities having a lower Cataract Surgical Coverage, thereby, underscoring the importance of eye care programs to address these inequalities.

## Introduction

Globally, 1.1 billion people have an untreated or preventable visual impairment (VI), including 43 million who are blind ^1, 2^. The major cause of blindness are cataract and uncorrected refractive error (URE) is the leading cause of moderate and severe VI ^2^. The burden of eye conditions is disproportionately higher in low- and middle-income countries with an estimated 39 million (91.8%) of the 43 million blind people living in these countries ^2^.

Gender disparities in blindness and VI have been noted with women accounting for 55% of blindness ^2, 3^. Studies across sub-Saharan Africa show that women, the elderly, people in rural areas, and those in the poorest households are at highest risk of cataract-related blindness and VI ^4^. Although factors like sex, economic status and disability have been identified as driving inequities, more data on the relationships between these variables is necessary ^5–8^.

Achieving universal eye health coverage is an integral step to achieving the Sustainable Development Goals, and Universal Health Coverage, as reducing the burden of VI will significantly improve individuals’ achievements in education, work, social participation and quality of life ^2^. The World Report on Vision describes the need for integrated person-centred eye care that adopts a health system perspective to address the significant eye health challenges facing many countries, recommending monitoring trends and evaluating progress as key actions for governments to meet their population eye health needs ^1^.

Nigeria, has a population of over 200 million people in 2022, with people 50 years and over accounting for about 10% ^9^. A national survey of VI in 2007 found the prevalence of blindness among adults aged over 40 years to be 4.2% (95%CI 3.8-4.6%) ^10^.

Kogi State, in northcentral Nigeria, is home to about 4 million people ^9^. In 2007, the prevalence of blindness in adults in this zone was 3.7% (95%CI 3.0-4.7%), prevalence of severe VI (SVI) was 1.7% (95%CI 1.2-2.5%) ^10^. An unpublished situational analysis report in 2018 estimated the cataract surgical rate (CSR) in Kogi to be below 100 per million population. There were four ophthalmologists and one government hospital and numerous private hospitals providing routine small incision cataract surgery with intraocular lens implantation.

No data on eye health status, needs, or service coverage had been collected in Kogi since the 2007 national survey. In 2019, the Kogi State Ministry of Health, supported by non-governmental organisation, Sightsavers, embarked on a new eye health programme with an aim to reach the most marginalised groups. A population-based blindness and VI survey was conducted at the outset to generate data to inform programme planning and baseline data to measure progress against. This paper reports the results of the survey.

## Methods

### Study design

A population-based cross-sectional survey, using a standardised rapid assessment of avoidable blindness (RAAB) methodology (described elsewhere), was conducted between September 2019 and March 2020 ^11^.

### Study population

This comprised adults aged 50 years and above resident in Kogi State for at least 6 months prior to the survey. Participants were recruited from 25^th^ September 2019 to 31^st^ March 2020.

### Sampling size and strategy

Sample size was calculated using the RAAB (version 5) software package. A minimum sample size of 4,150 was determined based on an estimated blindness prevalence of 3.75% ^10^, precision of 20% within a 95% confidence interval, non-compliance rate of 10% and design effect of 1.5.

A two-stage cluster sampling approach was adopted to select 4,150 eligible adults. First, 83 clusters were selected from a list of all villages (clusters) using probability proportionate to size methodology. Secondly, 50 eligible participants were enrolled from each village.

### Study procedures

The data collection teams comprised an ophthalmologist (team leader), an ophthalmic nurse and a cluster informer trained over five days by a certified RAAB trainer and passed an inter observer variation test according to RAAB guidelines before proceeding to the field.

Data were collected using an application on an android smartphone and uploaded to a cloud-based server. Precise location of villages (not households) was recorded using global positioning system (GPS) coordinates so that cluster level data could be visually mapped and geospatially analysed.

### Study protocols and definitions

All consenting participants underwent an eye examination conducted by the ophthalmologist following the standard RAAB protocol and answered additional questions regarding disability and household wealth . Using the WHO definitions, blindness was presenting VA (PVA) in the better eye less than 3/60, severe VI (SVI) was VA better than 3/60 to less than 6/60, moderate VI (MVI) was VA 6/60 to less than 6/18 ^12^. Refractive error was PVA<6/18 improving to 6/18 with pinhole. Cataract surgical coverage (CSC) is the number of operated persons with cataract as a proportion of those needing plus who have had surgery, effective CSC is the number of operated persons with cataract and have VA ≥6/18 as a proportion of those needing plus who have had surgery, and we used the new definitions presented in 2023 ^13^.

The eye examination recorded the following data: (i) PVA measurement of each eye (all participants). (ii) Pinhole VA of each eye if PVA<6/18. (iii) Lens examination of each eye with a torch in a darkened room (all participants). (iv) Posterior-segment examination with an ophthalmoscope of each eye PVA<6/18 where the principal cause cannot be attributed to refractive error, cataract, or corneal scarring. (v) Assessment of the major cause of VI of each eye PVA<6/18 and in persons where both eyes present VA<6/18 and the causes are not the same. (vi) Questions regarding where cataract surgery took place or why cataract surgery has not taken place where it is indicated.

Minor ocular conditions were treated by the team. Other conditions were referred to the nearest hospital.

Disability (reported elsewhere ^14^) was measured using the Washington Group Short Set (WGSS) on Functioning a tool comprising six questions regarding self-reported difficulties in six domains of functioning: seeing, hearing, climbing stairs or walking, remembering or concentrating, self-care, and communication ^15^. The severity of disability was measured on a graded scale - ‘no difficulty’, ‘some difficulty’, ‘a lot of difficulty’ and ‘cannot do at all’. Individuals reporting ‘a lot of difficulty’ or worse in at least one domain were considered to have a disability, creating a binary variable. Non-visual disability (a disability that excludes seeing difficulties) was defined as reporting ‘a lot of difficulty’ or worse in at least one domain excluding seeing.

The Nigeria Equity Tool (NET) ^16^, an internationally recognised tool designed to evaluate differences between social groups was used to categorise participants into one of five quintiles according to ownership of a list of household assets, with the poorest in the bottom quintile, and the wealthiest in the top quintile. The tool allows ascertaining the relative wealth of study participants compared to the national population. For the purposes of analysis, we created a binary variable by grouping the poorest three quintiles, and the wealthiest two.

### Data management and analysis

Data from the RAAB, Washington Group Short Set (WGSS) and Nigeria Equity Tool (NET) were captured on CommCare software deployed on Android-based smartphones. Each day, data were synced to the cloud, where it was downloaded and checked by a study team member. Data was analysed in two main stages. First, a standardised descriptive analysis was carried out using the RAAB software programme ^17^. Age- and sex-adjusted prevalence was calculated for all variables of blindness and VI. Second, Stata v15 was used to conduct chi-squared tests and adjusted logistic regression models to estimate the associations between eye health outcomes, sex, relative wealth and disability ^18^. Regression model standard errors were clustered to account for complex sample design. The level of statistical significance was calculated between variables at p<0.05.

### Ethics

The survey was approved by the Health Research Ethics Committee of Kogi State Ministry of Health Nigeria and satisfied all legal and regulatory requirements pertaining to research in Kogi State before commencement of the survey. Written consent was obtained from all participants and thumb prints substituted in non-literate participants, witnessed by an independent person – not a member of the study team – before participants were enrolled into the survey.

## Results

### Participant characteristics

The survey examined 3,926 adults (94.7% response rate) (Table 1); 2,283 (58.2%) were female and the sample had a higher proportion of men aged 80+ years than women; the oldest men were slightly older on average than the oldest women. From the sample, 212 (5.4%) had a non-visual disability (5.4% of men and 5.4% of women) and 1,078 (27.5%) adults lived in households belonging to the poorest quintile.

**Table 1:** Participant characteristics.

|  |  | Male |  | Female |  | Total |  |
| --- | --- | --- | --- | --- | --- | --- | --- |
|  |  | n | % | n | % | n | % |
| **Examination status of enrolled participants** |  |  |  |  |  |  |  |
|  | Examined | 1,643 | 94.8 | 2,283 | 95.2 | 3,926 | 94.7 |
|  | Not examined | 68 | 3.9 | 50 | 2.1 | 118 | 2.8 |
|  | Refused | 34 | 1.9 | 39 | 1.6 | 73 | 1.7 |
|  | Unable to communicate | 6 | 0.3 | 27 | 1.1 | 33 | 0.8 |
| **All examined** |  | 1,643 | 41.8 | 2,283 | 58.2 | 3,926 | 100.0 |
| **Age**<br>(years) | 50-59 | 747 | 45.5 | 1,091 | 47.8 | 1,838 | 46.8 |
|  | 60-69 | 502 | 30.6 | 634 | 27.8 | 1,136 | 28.9 |
|  | 70-79 | 232 | 14.0 | 339 | 14.8 | 571 | 14.6 |
|  | 80+ | 162 | 9.9 | 219 | 9.6 | 381 | 9.7 |
| **Disability**<br>(non-visual) | No | 1,554 | 94.6 | 2,160 | 94.6 | 3,714 | 94.6 |
|  | Yes | 89 | 5.4 | 123 | 5.4 | 212 | 5.4 |
| **Household wealth quintile** | Poorest | 488 | 29.7 | 590 | 25.8 | 1,078 | 27.5 |
|  | Q2 | 259 | 15.8 | 377 | 16.5 | 636 | 16.2 |
|  | Q3 | 295 | 18.0 | 506 | 22.2 | 801 | 20.4 |
|  | Q4 | 314 | 19.1 | 506 | 22.2 | 820 | 20.9 |
|  | Wealthiest | 287 | 17.5 | 304 | 13.3 | 591 | 15.1 |

### Prevalence of VI

Adjusted for age and sex, the all-cause prevalence of bilateral blindness was 3.6% (95% CI 3.0-4.2%), higher among men (4.1%, 95% CI 3.1-5.2%) than women (3.0%, 95% CI 2.4-3.9%), the difference not being statistically significant (Table 2). With best correction, the adjusted prevalence was 3.3% (95% CI 2.7-4.1%). Prevalence of presenting SVI was 2.0% (2.1% for men and 2.0% for women) and MVI 8.5% (8.8% for men and 8.1% for women).

**Table 2:**
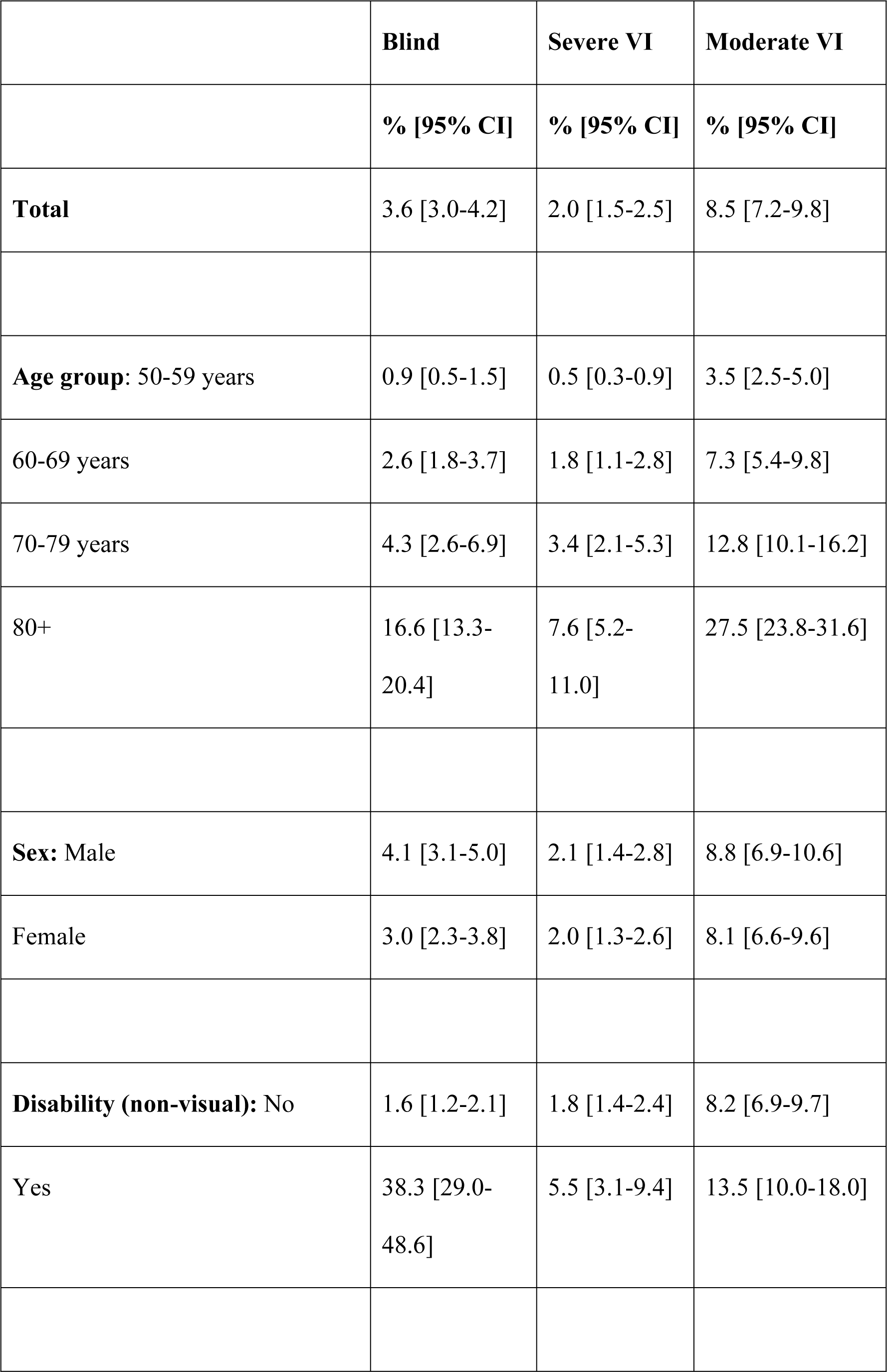
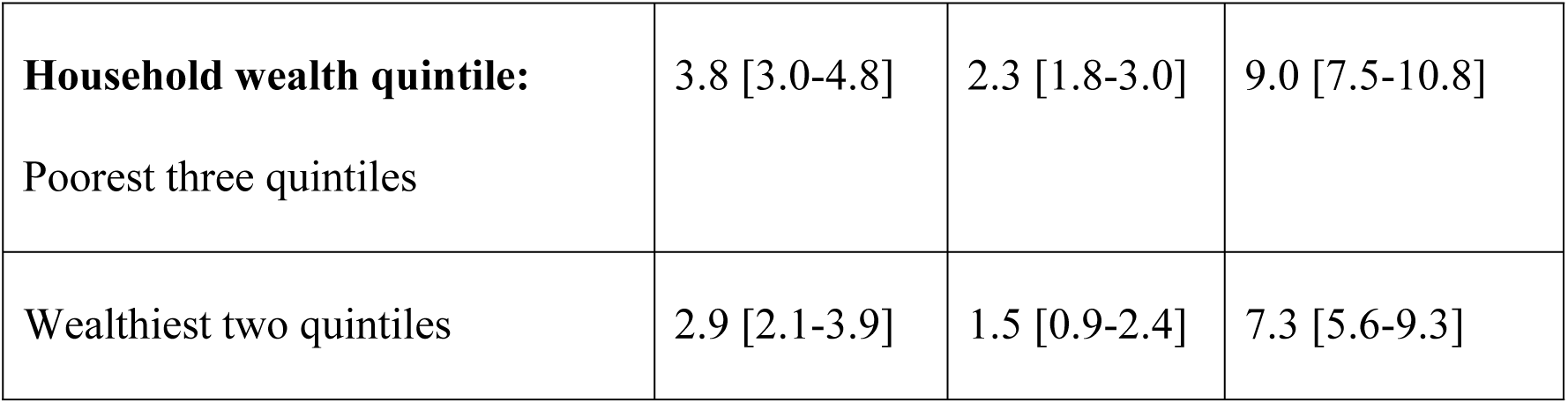
Age and sex adjusted prevalence of all cause bilateral presenting blindness, SVI and MVI (adjusted to population)

**Table 3:** Main causes of bilateral visual impairment.

|  | Blindness |  | Severe visual impairment |  | Moderate visual impairment |  |
| --- | --- | --- | --- | --- | --- | --- |
|  | n | % | n | % | n | % |
| Unoperated cataract | 73 | 55.3 | 52 | 67.5 | 118 | 36.2 |
| Glaucoma | 43 | 32.6 | 8 | 10.4 | 18 | 5.5 |
| Non-trachoma corneal opacity | 7 | 5.3 | 1 | 1.3 | 2 | 0.6 |
| Phthisis | 1 | 0.8 | 0 | - | 0 | - |
| Cataract surgical complications | 1 | 0.8 | 5 | 6.5 | 4 | 1.2 |
| Uncorrected refractive errors | 0 | - | 8 | 10.4 | 174 | 53.4 |
| Other | 7 | 5.3 | 3 | 3.9 | 10 | 3.1 |
| **Total** | **132** | **100.0** | **77** | **100.0** | **326** | **100.0** |

**Table 4:** Cataract surgical coverage and effective cataract surgical coverage (eyes) adjusted for age and sex (percentage)

|  | VA<3/60 | VA<6/60 | VA<6/18 |
| --- | --- | --- | --- |
| **Cataract surgical coverage (eyes)** |  |  |  |
| **Overall** | 48.0 | 36.7 | 22.5 |
| **Sex: Male** | 53.7 | 40.4 | 25.5 |
| Female | 40.3 | 31.5 | 18.2 |
| **Disability (non-visual): No** | 66.9 | 47.9 | 27.0 |
| Yes | 12.0 | 11.5 | 8.1 |
| **Household wealth: poorest three quintiles** | 39.4 | 27.6 | 16.5 |
| Wealthiest three quintiles | 61.2 | 53.9 | 35.0 |
| **Effective cataract surgical coverage (eyes)** |  |  |  |
| **Overall** | 31.0 | 24.2 | 14.8 |
| **Sex: Male** | 34.9 | 26.8 | 17.2 |
| Female | 25.5 | 20.4 | 11.4 |
| **Disability (non-visual): No** | 46.2 | 33.5 | 18.8 |
| Yes | 1.9 | 3.1 | 2.2 |
| **Household wealth: poorest three quintiles** | 24.3 | 17.3 | 10.6 |
| Wealthiest two quintiles | 41.1 | 37.0 | 23.6 |

**Table 5:** Barriers to cataract surgery.

|  | **Male** |  | **Female** |  | **Total** |  |
| --- | --- | --- | --- | --- | --- | --- |
|  | **n** | **%** | **n** | **%** | **n** | **%** |
| **Bilateral (BCVA<6/60) due to cataract** |  |  |  |  |  |  |
| Cost | 33 | 43.3 | 45 | 40.2 | 78 | 41.5 |
| Need not felt | 27 | 35.5 | 42 | 37.5 | 69 | 36.7 |
| Fear | 7 | 9.2 | 9 | 8.0 | 16 | 8.5 |
| Unaware treatment is possible | 4 | 5.3 | 3 | 2.7 | 7 | 3.7 |
| Cannot access treatment | 0 | 0.0 | 7 | 6.3 | 7 | 3.7 |
| Treatment denied by provider | 4 | 5.3 | 2 | 1.8 | 6 | 3.2 |
| Local reason | 1 | 1.3 | 4 | 3.6 | 5 | 2.7 |
| Total | 76 | 100.0 | 112 | 100.0 | 188 | 100.0 |

| **Unilateral (BCVA<6/60) due to cataract** |  |  |  |  |  |  |
| --- | --- | --- | --- | --- | --- | --- |
| Need not felt | 119 | 56.7 | 125 | 54.3 | 244 | 55.5 |
| Cost | 57 | 27.1 | 79 | 34.3 | 136 | 30.9 |
| Fear | 14 | 6.7 | 12 | 5.2 | 26 | 5.9 |
| Unaware treatment is possible | 11 | 5.2 | 2 | 0.9 | 13 | 3.0 |
| Local reason | 3 | 1.4 | 8 | 3.5 | 11 | 2.5 |
| Treatment denied by provider | 4 | 1.9 | 2 | 0.9 | 6 | 1.4 |
| Cannot access treatment | 2 | 1.0 | 2 | 0.9 | 4 | 0.9 |
| Total | 210 | 100.0 | 230 | 100.0 | 440 | 100.0 |

Adjusted for age and sex, the prevalence of all cause bilateral blindness among people living with additional, non-visual disabilities was 38.3% (95% CI 29.0-48.6%) compared to 1.6% (95%CI 1.2-2.1%; 𝜒^2^ = 771.9, p<0.001) among people without additional disabilities. Prevalence of SVI and MVI was estimated at 5.5% and 13.5%, respectively compared to 1.8 % and 8.2% among people without additional disabilities.

In general, bilateral blindness did not differ by household wealth quintile. However, the prevalence of VI at all levels was slightly higher among participants in the poorest three quintiles compared to the wealthiest two quintiles.

### Causes of VI

Cataract was the principal cause of bilateral blindness (73; 55.3% of all bilaterally blind people), glaucoma the second most important (43; 32.6%), and non-trachoma corneal opacities the third cause (7; 5.3%).

Cataract was the most important cause of SVI with 52 people (57.5% of all SVI cases), followed by glaucoma and uncorrected refractive errors affecting eight people each (10.4% each).

Uncorrected refractive error was the most important cause of MVI, affecting 174 people (53.4% of all MVI cases), followed by cataract (118; 36.2%) and glaucoma (18; 5.5%).

### Coverage of cataract services

Age and sex adjusted cataract surgical coverage (CSC) at the 3/60 level was 48.0%, higher among men than women (53.7% vs 40.3%). Among people with non-visual disabilities, CSC was 12.0% compared with 66.9% among people without non-visual disabilities. CSC was also higher among people living in households in the wealthiest two quintiles (41.1%) compared to the lowest three (24.3%).

Effective CSC at the 6/60 level was 31.0% overall, and higher among males (34.9%) than females (25.5%) and was extremely low among people with additional, non-visual disabilities (1.9%) compared to people with no additional disabilities (46.2%). It was also higher among people living in households belonging to the wealthiest two quintiles (41.1%) compared to the poorest three (24.3%).

Using presenting visual acuity (PVA), of the 116 eyes operated on for cataract, 61 (52.6%) had good outcome (VA>6/18) increasing to 71 (61.2%) with best correction; 16.4% were borderline, and 25.9% were (VA<6/60) even with pinhole correction (data not shown). Surgical complications were identified as the most common cause of the 36 poor visual outcomes in operated eyes (n=21, 58.3%), followed by selection (n=7) and sequelae (n=6). Both surgical complications (42.1%) and lack of spectacles (36.8%) were important causes of borderline outcomes.

### Barriers to cataract surgery

More than 40% of people with untreated bilateral cataracts cited cost as the main barrier to surgery; 36.7% did not feel they needed the operation; and 8.5% were afraid to have surgery. People with cataracts in one eye mostly described not feeling the need as the reason for not being operated (56.7%), followed by cost (27.1%). There was little difference between how men and women reported barriers to surgery.

## Discussion

The prevalence of blindness among persons 50 years and over was found to be 3.6%, almost unchanged from that reported in this part of Nigeria at 3.7% in 2007, suggesting that the point prevalence of blindness has not changed over the past 13 years ^10^. The prevalence of blindness was lower among females (3.0%, 95% CI 2.4-3.9) than males (4.1%, 95% CI 3.1-5.2) in this study, different from many other settings ^19, 20^.

We had expected a higher blindness prevalence as we are not aware of any past or current major blindness prevention initiatives in this state specifically targeting women. With an average CSR of 317 per million population per year in Nigeria ^21^, many people cannot access the few surgical sites in the state capital. Furthermore, the Nigerian population is aging and projected to reach 26 million in 2050 from 9 million reported in 2016 ^22^, the prevalence of blindness was expected to increase. Higher prevalence estimates were found in other parts of Nigeria and other Sub-Saharan African countries in more recent surveys: Plateau State 4.2% ^23^, Katsina State 5.3% ^20^, and lower levels in Sokoto State 1.9% ^19^.

As the major cause of blindness across Sub-Saharan Africa continues to be cataract ^2, 24^, our findings in Kogi State are consistent. Cataract surgical coverage for persons in our study was relatively lower (48.0%) than in other parts of Nigeria like Sokoto where in 2016 the CSC for persons was reported at 67.3% and shows that only half of people with an operable cataract had been operated on ^19, 25^. The Sokoto study did in fact show how in a ten-year period the CSC increased from 9% to 67% and prevalence of blindness reduced from 11.6% to 6.8% largely because of improved availability of human resources for cataract services. Availability of ophthalmologists as an indicator of the capacity of services meeting eye health needs has been shown to not be a strong predictor to the volume of cataract surgery and blindness prevalence elsewhere ^19, 26^.

It is possible that people travel to the neighbouring Kwara State, known to have good eye services, to have their surgery. Given that eye camps are intermittent, some people may stay long periods of time with only one eye operated on particularly considering the significant direct and indirect costs of a second eye surgery. Also, a person bilaterally blind from cataracts might not feel compelled to operate the other eye if he/she sees well with the other earlier operated eye. Conversely, if the initial surgery had a poor outcome, the individual may have a risk adverse attitude to future surgery.

Similarly, to other studies in sub-Saharan Africa, we found that women were more disadvantaged in accessing cataract services compared to men. The CSC for women in our study was 12% lower (42.1%) compared to men (54.9%). Gender inequalities in accessing cataract services are well known and have been documented in multiple settings ^6, 19, 27, 28^. In Nigeria, gender differences were documented in the Nigeria Blindness Survey, Plateau, and Sokoto states ^10, 19, 23^.

Effective CSC provides additional quantitative information on the proportion of operated individuals achieving good vision after surgery. In this study was found to be 31.0% overall, and significantly lower among women, poorer people, and those with additional, non-visual disabilities. Closing the gap in surgical coverage for females should be a deliberate attempt for all surgical service providers to ensure equity in attaining UHC.

The cataract surgical outcome (CSO) in this study identified the proportion of eyes with good VA following cataract surgery was 52.6% and increased to 61.2% with pinhole. The best corrected CSO is higher than what was recorded for the northcentral zone in the Nigeria Blindness Survey (54.9%) and slightly lower than in Sokoto State (69%) ^10,19^. The proportion of eyes with poor outcome following cataract surgery remains high in comparison with the Nigeria Blindness survey and may be attributed to surgical complications as recorded in this study. Quality surgical procedures such as biometry and appropriate intraocular lens power determination may not be available and could account for the leap in presenting versus best corrected visual outcome following surgery. In planning for more cataract surgeries, it is important to ensure quality and better outcomes.

This study identified cost as a major barrier to cataract surgery for those with bilateral cataracts. This is a significant challenge for patients as the average cost of cataract surgery per eye is 45,000 Naira ($64) whereas the minimum wage is 30,000 Naira ($42)^29^. Health insurance schemes have a low coverage with less than 5% of the working population enrolled in these schemes from 2005-2021, necessitating a high proportion of out-of-pocket payments for surgical services ^30^. For those with unilateral cataracts, no felt need was the biggest factor possibly because they still had unilateral functional vision. This is not different from the findings of other surveys in similar settings ^20, 31^.

### Strengths and Weaknesses

The survey response rate was high with 94.7%. Several reasons for this include the survey being conducted in the dry season when most farming activities were inactive, advance party information to the communities through the local cluster guides, and the survey team paying additional visits to absent enrolees. Women were more represented in the sample than men particularly in the younger age group 50-69 years. This may be related to the fact that there are marginally more women than men in the population within that age group, and this evens out in older ages. As such age- and sex-adjusted prevalence were reported. Also, this being partly a productive age group, it is possible that many men were out searching for jobs in the daytime.

As the RAAB tool is designed to identify avoidable blindness and coverage of cataract surgical services, it may have underestimated the burden of blindness and visual impairment due to posterior segment diseases such as glaucoma, diabetic retinopathy and age-related macular degeneration due to limitations in the diagnostic accuracy of the tool. Therefore, results must be interpreted with caution.

## Conclusions

This study has provided baseline information needed to set up an inclusive eye health program in Kogi State. The prevalence of blindness has remained relatively stable since the last survey in 2007 and the causes are mostly avoidable. The study has provided added information on the likely role that gender plays in the gaps in cataract surgical services and the burden of blindness among people with disabilities.

## Data Availability

All relevant data are contained in the manuscript. However, full data and permissions can be found on the RAAB Repository using the url below. Associated publications from the dataset are also linked to the url. https://www.raab.world/survey/nigeria-kogi-2020

## Acknowledgements

We acknowledge the members of the survey teams from Kogi State Ministry of Health and the Federal Medical Centre, Lokoja, the field assistants and participants who willingly consented to be examined.

